# Antibody responses to BNT162b2 mRNA vaccine: infection-naïve individuals with abdominal obesity warrant attention

**DOI:** 10.1101/2021.09.10.21262710

**Authors:** Alexis Elias Malavazos, Sara Basilico, Gianluca Iacobellis, Valentina Milani, Rosanna Cardani, Federico Boniardi, Carola Dubini, Ilaria Prandoni, Gloria Capitanio, Laura Valentina Renna, Sara Boveri, Matteo Carrara, Giovanni Spuria, Teresa Cuppone, Aurelia D’acquisto, Luca Carpinelli, Marta Sacchi, Lelio Morricone, Francesco Secchi, Elena Costa, Lorenzo Menicanti, Enzo Nisoli, Michele Carruba, Federico Ambrogi, Massimiliano Marco Corsi Romanelli

## Abstract

**Objective:** The excess of visceral adipose tissue might hinder and delay the immune response. How people with abdominal obesity will respond to mRNA vaccines against SARS-CoV-2 is yet to be established. We evaluated SARS-CoV-2-specific antibody responses after the first and second dose of the BNT162b2 mRNA vaccine comparing the response of individuals affected by abdominal obesity (AO) to those without, discerning between individuals with or without prior infection.

**Methods:** IgG neutralizing antibodies against the Trimeric complex (IgG-TrimericS) were measured at four time points: at baseline, at day 21 after vaccine dose-1, at one month and three months after dose-2. Nucleocapsid antibodies were assessed to detect prior SARS-CoV-2 infection. Waist circumference was measured to determine abdominal obesity.

**Results:** Between the first and third month after vaccine dose-2, the drop in IgG-TrimericS levels was more remarkable in individuals with AO compared to those without AO (2.44 fold [95%CI: 2.22-2.63] vs 1.82 fold [95%CI: 1.69-1.92], respectively, p<0.001). Multiple linear regression confirmed this result even when adjusting for possible confounders (p<0.001).

**Conclusions:** Our findings highlight the need to extend the duration of serological monitoring of antibody levels in infection-naive individuals with abdominal obesity, a higher-risk population category in terms of possible weaker antibody response.

## INTRODUCTION

Individuals with obesity and particularly those with predominant visceral adipose tissue (VAT) accumulation are at significant risk of developing a more severe coronavirus disease 2019 (COVID-19) [1–3].

Messenger RNA (mRNA) vaccines against severe acute respiratory syndrome coronavirus-2 (SARS-CoV-2), the causative agent of COVID-19, hold great promise for curbing the spread of infection and accelerating the timeline towards a possible level of herd immunity [4]. Nevertheless, researchers fear that vaccines against SARS-CoV-2 might not be as effective in people with obesity [5], since studies on vaccines against influenza, hepatitis B, and rabies have shown a reduced immune response in these individuals [5,6]. To date, the available clinical evidence demonstrates that the efficacy of mRNA vaccines against SARS-CoV-2 does not differ among individuals with obesity compared to those without obesity [4,7].

However, these analyses have focused on the definition of obesity assessed through body mass index (BMI), yet this is not the best indicator of adiposity since it does not take into account the amount and distribution of body fat, which can differ broadly among people with the same BMI [8]. The excess of VAT is considered the main culprit in inflammatory diseases linked to obesity and it is an indicator of increased ectopic fat, which might hinder and delay the immune response, as highlighted by COVID-19 [1,2,5,8–11]. How people with abdominal obesity (AO) will respond to mRNA vaccines against SARS-CoV-2 is yet to be established. To this end, we evaluated SARS-CoV-2-specific antibody responses after the first and second dose of the BNT162b2 mRNA vaccine in a large cohort of healthcare workers. We compared the response of individuals affected by AO to those without AO, discerning between individuals with, or without prior SARS-CoV-2 infection.

## METHODS

### Study design and population

VARCO-19 study is an ongoing observational prospective cohort study started in January 2021 and lasting until March 2022. All participants provided written informed consent. The protocol was approved by IRCCS Policlinico San Donato Internal Scientific Committee – VARCO-19 Protocol January 2021. (Project code RC 5.17.01) and by National Ethics Committee of INMI IRCCS Lazzaro Spallanzani (Opinion No. 403 of the 2020/2021 Trial Registry). The vaccination itself was not part of the study.

We collected blood samples from a cohort of healthcare workers who received two doses of BNT162b2 mRNA vaccine at IRCCS Policlinico San Donato, a large academic medical centre in Milan, Italy. One thousand and sixty patients met the following inclusion criteria: (1) age >18 years; (2) two doses of BNT162b2 mRNA vaccine. Pregnancy was an exclusion criterion. All participants provided data on medical history, pharmacotherapy (if any), smoking status and symptoms if reported prior SARS-CoV-2 infection.

### Anthropometric parameters

Anthropometric parameters were assessed at baseline. Waist circumference was measured midway between the lower rib and the iliac crest to the closest 1.0 cm. The cut-off to determine AO was 94 cm in men and 80 cm in women [8, 12]. BMI was calculated as weight (kilograms) divided by height (meters) square [13].

### Serological testing

Antibody levels were measured at four time points: at baseline, at day 21 after vaccine dose-1, at one month (within 30-40 days) and three months (within 90-100 days) after dose-2.

Serological testing for SARS-CoV-2 glycoprotein-specific anti-spike (S) IgG was performed using LIAISON® assay, which measures IgG neutralizing antibodies against the Trimeric complex (IgG-TrimericS), which includes the receptor-binding-domain (RBD) and N-terminal-domain (NTD) sites from the three S1 subunits. We used nucleocapsid (N) antibodies to detect prior infection. Given that the BNT162b2 vaccine delivers mRNA encoding only for S-protein, the expected elicited response is the production of IgG-TrimericS levels and not N-antibodies, which represent a durable marker and indicator of post-infectious status [14–16]. Antibody assay methods are discussed in more detail in the supporting information. Results were reported as Binding Antibody Unit (BAU)/ml (≥33.8 BAU/ml is positive).

### Statistical analyses

We expressed antibody levels as geometric mean (standard error, SE). Confidence intervals of the geometric means were calculated using log transformed data and t-tables with back-transformation. Individuals were categorized according to AO and BMI-classes with and without prior SARS-CoV-2 infection. For comparing between-group continuous values, the nonparametric Kruskal-Wallis test was used. We used multivariable linear regression to account for possible confounding and evaluate the three months difference in absolute variation of titre levels starting from one month after dose 2 in individuals with and without AO (or BMI-classes). The model was also adjusted for one month after dose 2 antibody levels, gender, age, smoke, prior SARS-CoV-2 infection and the interaction between prior infection and AO (or BMI-classes). The null hypothesis will be refused with p<0.05. All statistical analyses were done with SAS version 9.4 (SAS Institute, Cary, NC).

## RESULTS

Baseline characteristics of patients are reported in Table 1. Vaccine recipients (n=1060), who provided at least three blood samples for antibody testing, were aged 41.4±12.9 years, 62% were female, and 93% were Caucasian: 825 vaccine recipients (186 with prior infection) provided blood samples once after dose-1 and twice after dose-2; 235 vaccine recipients (54 with prior infection) also provided baseline (pre-vaccine) samples.

**Table 1.** Demographic and clinical characteristics of individuals with or without abdominal obesity and with or without prior SARS-CoV-2 infection.

|  |  | Abdominal Obesity (n=492) |  |  | No abdominal obesity (n=568) |  |  |
| --- | --- | --- | --- | --- | --- | --- | --- |
|  | Total<br>(n=1060) | No Prior SARS-CoV-2<br>infection (n=380) | Prior SARS-CoV-2<br>infection (n=112) | p-value | No Prior SARS-CoV-2<br>infection (n=440) | Prior SARS-CoV-2<br>infection (n=128) | p-value |
| Age, years | 41.42±12.95 | 47.50±12.25 | 44.54±10.99 | 0.022 | 36.75±11.88 | 36.67±11.80 | 0.949 |
| Race |  |  |  | 0.001* |  |  | 0.356* |
| Caucasian | 991 (93.49%) | 361 (95.00%) | 94 (83.93%) |  | 416 (94.55%) | 120 (93.75%) |  |
| Latin-American | 44 (4.15%) | 12 (3.16%) | 11 (9.82%) |  | 15 (3.41%) | 6 (4.69%) |  |
| Asia | 1 (0.09%) | - | - |  | 0 (0.00%) | 1 (0.78%) |  |
| African | 9 (0.85%) | 2 (0.53%) | 3 (2.68%) |  | 4 (0.91%) | 0 (0.00%) |  |
| Arabic | 15 (1.42%) | 5 (1.32%) | 4 (3.57%) |  | 5 (1.14%) | 1 (0.78%) |  |
| Gender |  |  |  | 0.058 |  |  | 0.407 |
| Male | 400 (37.74%) | 153 (40.26%) | 34 (30.36%) |  | 161 (36.59%) | 52 (40.63%) |  |
| Female | 660 (62.26%) | 227 (59.74%) | 78 (69.64%) |  | 279 (63.41%) | 76 (59.38%) |  |
| Smoking status |  |  |  | 0.059 |  |  | 0.360 |
| Smoker | 167 (15.75%) | 69 (18.16%) | 10 (8.93%) |  | 73 (16.59%) | 15 (11.72%) |  |
| Former smoker | 25 (2.36%) | 12 (3.16%) | 3 (2.68%) |  | 7 (1.59%) | 3 (2.34%) |  |
| Non smoker | 868 (81.89%) | 299 (78.68%) | 99 (88.39%) |  | 360 (81.82%) | 110 (85.94%) |  |
| Comorbidities |  |  |  |  |  |  |  |
| Hypertension | 97 (9.15%) | 67 (17.63%) | 15 (13.39%) | 0.290 | 13 (2.95%) | 2 (1.56%) | 0.387 |
| Diabetes mellitus | 15 (1.42%) | 8 (2.11%) | 6 (5.36%) | 0.099* | 1 (0.23%) | 0 (0.00%) | 1.00* |
| Cardiovascular diseases | 32 (3.02%) | 19 (5.00%) | 3 (2.68%) | 0.296 | 9 (2.05%) | 1 (0.78%) | 0.338 |
| Dyslipidemia | 44 (5.15%) | 26 (6.84%) | 7 (6.25%) | 0.826 | 8 (1.82%) | 3 (2.34%) | 0.704* |
| Cancer | 4 (0.38%) | 4 (1.05%) | 0 (0.00%) | 0.579 | 0 (0.00%) | 0 (0.00%) | - |
| Other | 73 (6.89%) | 34 (8.95%) | 6 (5.36%) | 0.222 | 21 (4.77%) | 12 (9.38%) | 0.050 |
| Anthropometric measurements |  |  |  |  |  |  |  |
| Weight, kg | 70.40±15.01 | 78.32±14.46 | 80.66±15.92 | 0.143 | 63.06±10.39 | 64.17±11.85 | 0.918 |
| Height, cm | 168.35±9.05 | 168.46±9.18 | 167.06±8.75 | 0.151 | 168.55±8.74 | 168.47±9.95 | 0.922 |
| Waist, cm | 85.30±13.55 | 94.95±11.12 | 96.80±12.54 | 0.135 | 76.52±7.90 | 76.77±8.55 | 0.753 |
| Waist male, cm | 93.22±12.11 | 103.02±8.71 | 103.90±11.29 | 0.618 | 84.36±5.99 | 84.87±6.47 | 0.602 |
| Waist female, cm | 80.50±12.05 | 89.52±9.09 | 93.71±11.84 | 0.001 | 72.00±4.67 | 71.24±4.36 | 0.203 |
| WHtR | 0.51±0.08 | 0.56±0.06 | 0.58±0.07 | 0.018 | 0.45±0.04 | 0.46±0.04 | 0.718 |
| BMI, kg/m<sup>2</sup> | 24.75±4.41 | 27.50±3.96 | 28.81±4.74 | 0.003 | 22.10±2.44 | 22.09±2.46 | 0.983 |
| BMI classes |  |  |  | 0.065 |  |  | 0.257 |
| Underweight | 37 (3.49%) | - | - |  | 32 (7.27%) | 5 (3.91%) |  |
| Normal weight | 594 (56.04%) | 106 (27.89%) | 22 (19.64%) |  | 361 (82.05%) | 105 (82.03%) |  |
| Overweight | 311 (29.34%) | 191 (50.26%) | 55 (49.11%) |  | 47 (10.68%) | 18 (14.06%) |  |
| Obesity | 118 (11.13%) | 83 (21.84%) | 35 (31.25%) |  | - | - |  |
| Prior SARS-CoV-2 infection symptoms |  |  |  |  |  |  |  |
| Mild or asymptomatic | 120 (50.0%) | - | 52 (46.85%) | - | - | 68 (53.54%) | - |
| Symptomatic | 118 (49.12%) | - | 59 (53.15%) | - | - | 59 (46.46%) | - |
| IgG-TrimericS antibody levels |  |  |  |  |  |  |  |
| Baseline<sup>£</sup> | 10.7 [8.9-13.0] | 5.2 [4.7-5.8] | 164.1 [74.2-363.0] | <0.001 | 6.0 [5.3-6.9] | 53.4 [39.9-88.4] | <0.001 |
| 21 day after dose 1<sup>§</sup> | 536.4 [501.7-573.46] | 270.4 [243.4-300.4] | 1910.6 [1768.1-2064.6] | <0.001 | 484.6 [449.2-522.7] | 1912.2 [1805.4-2025.2] | <0.001 |
| 1 month (within 30-40 days) after dose 2<sup>§</sup> | 1693.4 [1656.7-1730.9] | 1434.6 [1365.7-1507.0] | 1992.4 [1937.1-2049.2] | <0.001 | 1780.0 [1739.7-1821.2] | 2024.2 [1977.2-2072.2] | <0.001 |
| 3 months (within 90-100 days) after dose 2<sup>§</sup> | 929.7 [889.75-971.51] | 591.5 [548.1-638.3] | 1691.5 [1564.3-1829.2] | <0.001 | 983.1 [931.5-1037.6] | 1740.5 [1646.5-1840.0] | <0.001 |

Abdominal obesity (waist circumference $\geq 94$ cm for men, $\geq 80$ cm for women); No abdominal obesity (waist circumference $< 94$ cm for men, $< 80$ cm for women); Underweight (BMI $< 18.5$ kg/m<sup>2</sup>); Normal weight (BMI from 18.5 to $< 25$ kg/m<sup>2</sup>); Overweight (BMI from 25 to $< 30$ kg/m<sup>2</sup>); Obesity (BMI $\geq 30$ kg/m<sup>2</sup>). Body Mass Index (BMI); Waist-to-height ratio (WHtR); Antibody levels are expressed as BAU/ml (BAU = Binding Antibody Units) and are presented as geometric mean [confidence interval].
Data are n (%), mean (SD). To compare variables between different groups, the Chi-square or \*Fisher's exact test was used for categorical variables. T-test or Wilcoxon rank sum test is used to compare unpaired means in normally or non-normally continuous distributed variables, respectively.
<sup>§</sup>n total=1060; <sup>£</sup>n total=235

At baseline, among infection-naïve individuals, as expected, there was no difference in IgG-TrimericS levels between individuals with or without AO (figure-A). At other times, among infection-naïve individuals, IgG-TrimericS levels were significantly lower in individuals with AO than in those without AO, (figure-B). Two aspects are noteworthy: (i) at one month after dose-2, individuals with AO reached a lower peak of IgG-TrimericS levels than individuals without AO (geometric mean BAU/ml [confidence interval], 1434.6 BAU/ml [95%CI: 1365.7–1507.0] vs 1780.0 BAU/ml [95%CI: 1739.7–1821.2], p<0.001), (figure-B); (ii) between the first and third month after vaccine dose-2, the drop in IgG-TrimericS levels was more remarkable in individuals with AO compared to those without AO (2.44 fold [95%CI: 2.22–2.63] vs 1.82 fold [95%CI: 1.69–1.92], respectively, p<0.001) (figure-B).

**Figure.**
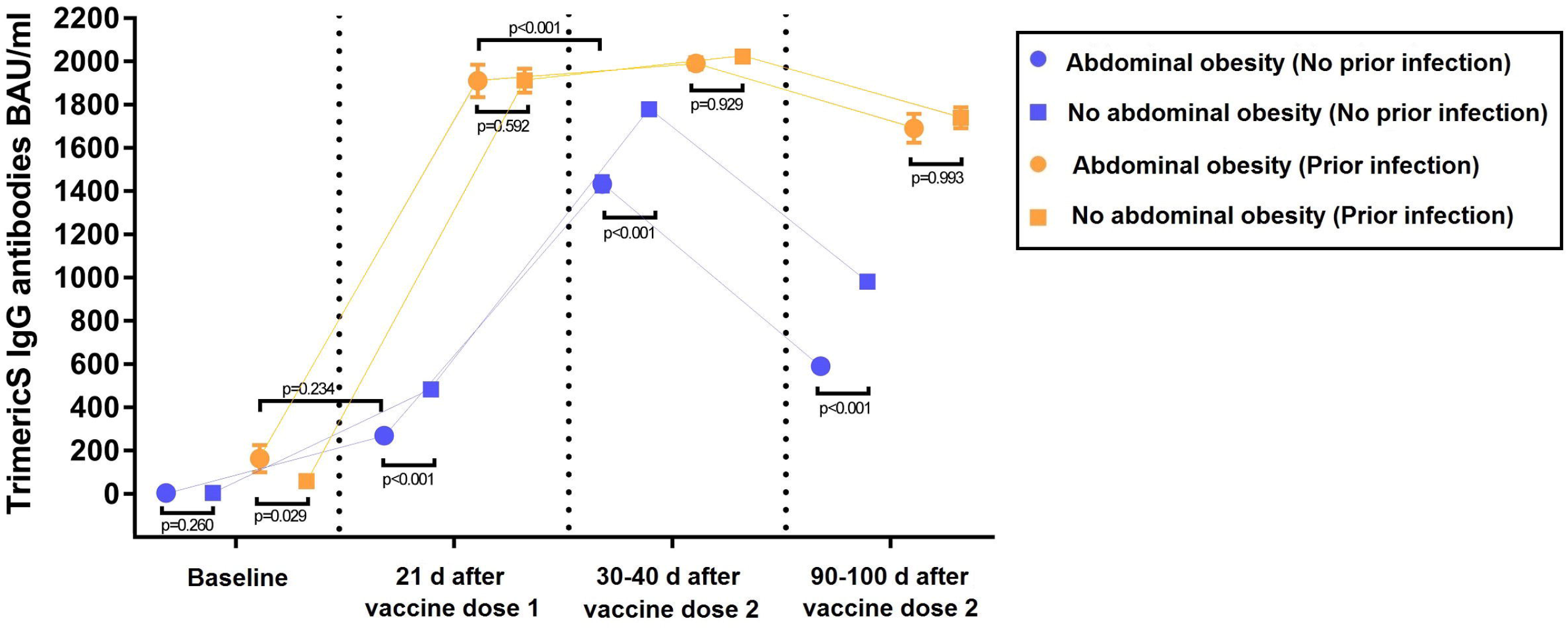
IgG-TrimericS antibody response to mRNA SARS-CoV-2 vaccination in individuals with or without abdominal obesity, discerning between individuals with or without prior SARS-CoV-2 infection. Abdominal obesity (waist circumference ≥94 cm for men, ≥80cm for women); no abdominal obesity (waist circumference <94cm for men, <80cm for women). Antibody levels are expressed as BAU/ml (BAU = Binding Antibody Units) and are presented as geometric mean [confidence interval]. (A) Infection-naïve individuals with abdominal obesity (n=79) and those without abdominal obesity (n=102) both at baseline (5.2 BAU/ml [95%CI: 4.7–5.8] *vs* 6.0 BAU/ml [95%CI: 5.3–6.9], p=0.26). (B) Infection-naïve individuals with abdominal obesity (n=380) and those without abdominal obesity (n=440): both at 21 days after dose-1 (270.4 BAU/ml [95%CI: 243.4–300.4] *vs* 484.6 BAU/ml [449.2–522.7], p<0.001), both at 1 month (within 30-40 days) after dose-2 (peak) (1434.6 BAU/ml [95%CI: 95%CI: 1365.7–1507.0] vs 1780.0 BAU/ml [95%CI: 1739.7–1821.2], p<0.001), both at 3 months (within 90-100 days) after dose-2 (591.5 BAU/ml [95%CI: 548.1–638.3] *vs* 983.1 BAU/ml [95%CI: 931.5–1037.6], p<0.001). (C) Prior infection individuals with abdominal obesity (n=23) and those without abdominal obesity (n=31) at both baseline (164.1 BAU/ml [95%CI: 74.2–363.0] *vs* 59.4 BAU/ml [95%CI: 39.9–88.4], p=0.029). (D) Prior infection individuals with abdominal obesity (n=112) and those without abdominal obesity (n=128): both at 21 days after dose-1 (1910.6 BAU/ml [95%CI: 1768.1–2064.6] *vs* 1912.2 BAU/ml [95%CI: 1805.4–2025.2], p=0.592), both at 1 month (within 30-40 days) after dose-2 (1992.3 BAU/ml [95%CI: 1937.1–2049.2] *vs* 2024.1 BAU/ml [95%CI: 1977.2–2072.2], p=0.929), both at 3 months (within 90-100 days) after dose-2 (1691.5 BAU/ml [95%CI: 1564.3–1829.2] *vs* 1740.5 BAU/ml [95%CI: 1646.5–1840.0], p=0.993). (E) Individuals with prior infection at baseline (n=23) and infection-naïve individuals with abdominal obesity at 21 days after dose 1 (n=380) (164.1 BAU/ml [95%CI: 74.2–363.0] *vs* 270.4 BAU/ml [95%CI: 243.4–300.4], p=0.234). (F) Infection-naïve individuals with abdominal obesity (n=380) at 1 month (within 30-40 days) after dose-2 (peak) and those with prior infection at 21 days after dose-1 (peak) (n=112) (1434.6 BAU/ml [95%CI: 1365.7–1507.0] *vs* 1910.6 BAU/ml [95%CI: 1768.1–2064.6], p<0.001).

At baseline, among individuals with prior infection those with AO had higher IgG-TrimericS levels than those without AO (164.1 BAU/ml [95%CI: 74.2–363.0] vs 59.4 BAU/ml [95%CI: 39.9–88.4], p=0.029) (figure-C). There was no difference between the two groups at other times (figure-D).

Among individuals with AO, IgG-TrimericS levels were slightly lower in individuals with prior infection at baseline than in infection-naïve individuals after dose-1 (164.1 BAU/ml [95%CI: 74.2–363.0] vs 270.4 BAU/ml [95%CI: 243.4–300.4], p=0.234), (figure-E).

Among individuals with AO, IgG-TrimericS levels were significantly lower in infection-naïve individuals after dose 2 than in previously infected individuals after dose-1 (1434.6 BAU/ml [95%CI: 1365.7–1507.0] vs 1910.6 BAU/ml [95%CI: 1768.1–2064.6], p<0.001) (figure-F).

Analysis of these data by multivariable linear regression showed evidence of interaction between AO and SARS-CoV-2 infection (p=0.002) (table 2). Specifically, AO is associated with a drop in absolute IgG-TrimericS levels in infection-naïve individuals at three months after dose 2, regardless of gender, age, or smoking, and IgG-TrimericS levels reached at 1 month after dose-2 (159.2 BAU/ml [95%CI: 96.9–221.4], p<0.001). No interaction was evinced using BMI-classes in the same regression model (p=0.267) (table 2).

**Table 2.**
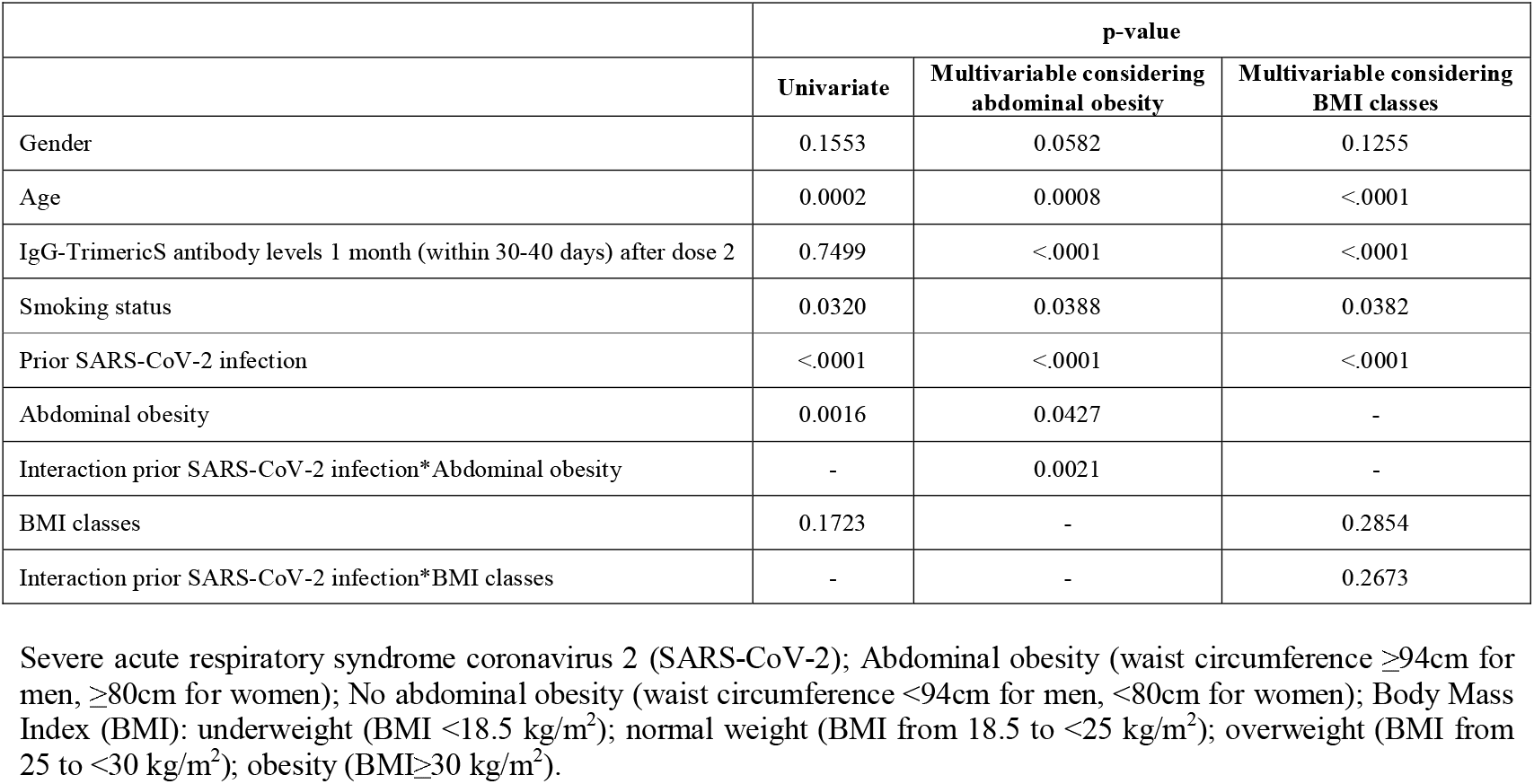
Univariate and multivariable linear regression to evaluate the three months (within 90-100 days) difference in absolute variation of IgG-TrimericS antibody levels starting from 1 month after dose 2.

|  | p-value |  |  |
| --- | --- | --- | --- |
|  | Univariate | Multivariable considering abdominal obesity | Multivariable considering BMI classes |
| Gender | 0.1553 | 0.0582 | 0.1255 |
| Age | 0.0002 | 0.0008 | <.0001 |
| IgG-TrimericS antibody levels 1 month (within 30-40 days) after dose 2 | 0.7499 | <.0001 | <.0001 |
| Smoking status | 0.0320 | 0.0388 | 0.0382 |
| Prior SARS-CoV-2 infection | <.0001 | <.0001 | <.0001 |
| Abdominal obesity | 0.0016 | 0.0427 | - |
| Interaction prior SARS-CoV-2 infection*Abdominal obesity | - | 0.0021 | - |
| BMI classes | 0.1723 | - | 0.2854 |
| Interaction prior SARS-CoV-2 infection*BMI classes | - | - | 0.2673 |

Severe acute respiratory syndrome coronavirus 2 (SARS-CoV-2); Abdominal obesity (waist circumference $\geq 94$ cm for men, $\geq 80$ cm for women); No abdominal obesity (waist circumference $< 94$ cm for men, $< 80$ cm for women); Body Mass Index (BMI): underweight (BMI $< 18.5$ kg/m<sup>2</sup>); normal weight (BMI from 18.5 to $< 25$ kg/m<sup>2</sup>); overweight (BMI from 25 to $< 30$ kg/m<sup>2</sup>); obesity (BMI $\geq 30$ kg/m<sup>2</sup>).

## DISCUSSION

Our results showed that infection-naïve individuals with AO had lower antibody development over time compared to individuals without AO. They reached a lower antibody peak and they had a more significant drop in antibody levels at three months after dose-2, regardless of possible confounders.

It is worth noting that infection-naïve individuals with AO reached a lower antibody peak after dose-2 than individuals with AO and prior infection after a single vaccine dose. Recent studies showed that individuals with previous SARS-CoV-2 infection may have a superior humoral response after the first dose of BNT162b2 compared with uninfected individuals who were fully vaccinated with two doses of BNT162b2 [17, 18]. Furthermore, we have found that among individuals with prior infection those with AO had higher IgG-TrimericS levels than those without AO at baseline. These data are consistent with a recent paper in which it was observed that individuals with severe obesity and prior infection exhibited a higher neutralising antibody titre likely due to more severe COVID-19 [11].

A recent small study, conducted over a period of only four weeks after vaccine dose-2, hypothesised that central obesity is associated with lower antibody titres following mRNA vaccines against SARS-CoV-2 [19]. The lower antibody response and its more significant drop over time, highlighted by classifying the population by AO phenotype rather than BMI-based obesity, can be explained in several ways, the most important being the greater chronic-inflammatory status due to an excess of abdominal VAT, which could compromise the immune system [1,5,6].

Remarkably, the available clinical evidence and a recent position statement showed that vaccines are not less effective for individuals with obesity and that they are an important protection against COVID-19 [4,7]. Antibody titre following SARS-CoV-2 vaccination cannot predict alone the risk of developing COVID-19, and even low antibody levels could be equally protective against infection. Moreover, in a recent pre-print study, it was shown that BNT162b2 mRNA vaccine efficacy tends to gradually decrease after six months, while maintaining a high safety profile [20].

Limitations of our study include lack of measurement of virus-specific T-cell. Anti-N antibodies were assessed only once to determine a prior SARS-CoV-2 infection. In addition, we did not evaluate pro-inflammatory markers of inflammation.

Hence, our findings highlight the need to extend the duration of serological monitoring of antibody levels in infection-naïve individuals with AO, a higher risk population category in terms of possible weaker antibody response.

## Supporting information

Supporting informations

## Data Availability

The data that support the findings of this study are available from the corresponding author [A.E.M] upon reasonable request.

## AKNOLEDGEMENT

We acknowledge the samples collection effort of Matteo Maiocchi, Michele Iovane, Mirco Bonaccorso, Sofia Sichel from Endocrinology Unit, Clinical Nutrition and Cardiovascular Prevention Service at IRCCS Policlinico San Donato, San Donato Milanese, Milan, Italy, Ketty Miani, Ganiyat Adenike Ralitsa Adebanjo and Mariapia Zagaria from residency program in Clinical Pathology and Clinical Biochemistry at University of Milano, Milan, Italy and of Maria Luisa Trovato from Operative Unit of Laboratory Medicine1-Clinical Pathology, Department of Pathology and Laboratory Medicine, IRCCS Policlinico San Donato, San Donato Milanese, Milan, Italy.

