## Supporting informations for "Antibody responses to BNT162b2 mRNA vaccine: infection-naïve individuals with abdominal obesity warrant attention"

### SUPPORTING INFORMATION

#### Serological testing

Serological testing for SARS-CoV-2 anti-spike (S) antibodies was performed using the LIAISON® XL Analyzer by DiaSorin (Italy), a fully automated chemiluminescence analyzer (CLIA). We used LIAISON® SARS-CoV-2 TrimericS IgG assay that measures IgG antibodies against the Trimeric complex, including the receptor binding domain (RBD) and N-terminal domain (NTD) sites from the three subunit S1. Trimeric spike glycoprotein is the stabilized native form of the SARS-CoV-2 Spike protein, which may elicit an accurate detection of IgG Neutralizing antibodies, consisting of two subunits S1 and S2 [1]. It has been demonstrated that Trimeric Spike protein used in this test can detect the complete variety of the natural immune response to the virus, instead of limiting the detection to antibodies against single epitopes such as RBD, commonly used by other assays [2]. This allows detecting a broader population of antibodies generated by the body's immune response, reducing the risk of false negative results. The quantitative measure of IgG antibody goes between a range of 4.81 and 2080 BAU/mL (BAU = Binding Antibody Units).

Antibody levels were measured at four time points: at baseline (pre-vaccine), at days 21 after vaccine dose 1 and within 30-40 days and 90-100 after dose 2. Results are reported as negative (<33.8 BAU/ml) or positive (≥33.8 BAU/ml).

Qualitative assessment of SARS-CoV-2 anti-nucleocapsid (anti-N) antibodies was performed using the Cobas E 601 module by Roche (Germany) that exploits electrochemiluminescence (ECL) technology. We used Elecsys Anti-SARS-CoV-2, a sandwich test that uses a recombinant protein representing the nucleocapsid antigen to determine antibodies against SARS-CoV-2. Results are reported qualitatively with a cutoff index (COI) of <1.0 interpreted as nonreactive (COVID-19–) and ≥1.0 interpreted as reactive (COVID-19+). Clinical sensitivity is 98.8%, and specificity is 99.9%. A previous SARS-CoV-2 infection was determined by positive detection of anti-nucleocapsid antibodies (COVID-19+).

Accordingly, we determined prior infection status based on a concordance of the presence of N-antibodies, data documented in health records and self-reported survey information collected.

In addition, as per hospital policy, all participants had undergone monthly repeated nasopharyngeal swab for detection of SARS-CoV-2 RNA through a Reverse Transcription and Real-Time Polymerase Chain Reaction, using GeneFinder™ COVID-19 Plus RealAmp kit (OSANG Healthcare). Before vaccine dose-2, three individuals of our sample, 2 females and 1 male, non-smokers, one with abdominal obesity, resulted positive to nasopharyngeal swab to detect reverse-transcriptase polymerase chain reaction (RT-PCR) to SARS-CoV-2 after vaccine dose-1. Two individuals were asymptomatic and the one with abdominal obesity had mild symptoms like fever and cold. They were excluded from further analysis.
